# Promoting psychological health and overall wellness in female Veterans with military sexual trauma through complementary health Interventions: A pilot study

**DOI:** 10.1101/2024.06.11.24308617

**Authors:** Shengnan Sun, Akiva Singer, Nithya Ganesh, Ann Feder, Lauren Byma, Lisa Wang, Monique McClenton, Hanga Galfalvy, Fatemeh Haghighi

## Abstract

**Introduction:** Military Sexual Trauma (MST) has been associated with long-term negative outcomes such as increased rates of cardiovascular disease, PTSD, and suicidal thoughts and behaviors. While evidence supports the effectiveness of psychotherapeutic approaches as treatments for MST and related PTSD symptoms, these interventions have limited impact, attributed to perceived stigma with high dropout rates in female Veterans. Complementary and integrative health (CIH) interventions provide an alternative that may be more acceptable and can help transition Veterans into mental health treatments. Although research has found individual CIH interventions to be both effective and acceptable treatments for MST-related PTSD amongst female Veterans, lacking are evaluations of interventions that combine multiple CIH modalities or specifically in populations of at-risk female Veterans with history of suicidal ideation or behavior. Thus, this quality improvement project aimed to assess the impact of a multimodal CIH intervention on mental and physical health symptoms specifically in female Veterans with MST.

**Materials and Methods:** Female Veterans (N=38) with and without a history of MST participated in an ongoing multimodal CIH intervention. Programming took place over multiple 4-week long cohorts during which Veterans engaged in meditation and mindfulness, physical exercise, nutrition, and motivational curricula. Mental health symptoms and other factors related to suicide risk were assessed before and after program participation, using measures including the PTSD Checklist for DSM-5 (PCL), Perceived Stress Scale (PSS), Beck Anxiety Scale (BAS), Beck Depression Inventory II (BDI), Patient Health Questionnaire (PHQ-9), Measure of Current Status (MoCS), Pittsburgh Sleep Quality Inventory (PSQI), and the Defense and Veterans Pain Rating Scale (DVPRS). These measures were assessed for baseline differences between those with vs. without MST, pre-post intervention differences, and with ANCOVA for pre-post group differences accounting for baseline score.

**Results:** Most participants reported a history of MST (68%), with those endorsing MST also having significantly worse baseline scores for depressive symptoms (PHQ-9 and BDI; p = 0.0402 and 0.0360 corresponding), PTSD symptoms (PCL; p =0.0360), perceived stress (PSS; p = 0.0254), and sleep quality (PSQI; p= 0.0121) than those without MST. No significant baseline differences were found for hopelessness (BHS), perceived coping ability (MoCS), anxiety (BAS), and pain (DVPRS). Significantly greater improvement in depressive symptoms (PHQ-9 and BDI; p = 0.0156, p = 0.0211), and perceived stress (PSS; p = 0.0351) was found in comparison of the MST vs. no MST group but not for the other scales; findings were no longer significant after accounting for baseline group differences across these measures. Finally, in the subset of Veterans with histories of suicide ideation or attempt, medium to large treatment effects were found for all measured outcomes (Cohen’s |d| > .54).

**Conclusions:** The results of this quality improvement project add to the growing body of evidence demonstrating that CIH interventions can be effective in attenuating mental health symptoms related MST and extend these findings to female Veteran populations at-risk for suicide. Additionally, they lend initial weight to the ability of treatments integrating multiple CIH interventions to have similar positive impacts to the singular interventions typically studied.

## INTRODUCTION

The Department of Veterans Affairs (VA) defines Military Sexual Trauma (MST) as any sexual activity that occurs during active military service against one’s will^1^. Although MST impacts both males and females, the latter are disproportionately affected, with 38.4% of female Veterans and Service Members reporting MST experience during service compared to 3.9% of their male counterparts^2^. MST has a long-term negative impact on both medical as well as mental health outcomes, with increased rates of obesity and cardiovascular disease amongst females with MST and depression and PTSD^3^. Of relevance to the present study, MST has also been associated with an increased risk of suicidal behavior and suicidal ideation, with 75% experiencing ideation and roughly 41% attempting suicide^4^. Evidence-based psychotherapies including cognitive processing therapy (CPT) and prolonged exposure therapy (PE) are two common therapeutic treatments which emphasize approaching memories and changing beliefs that are associated with experienced trauma and have been applied in treatment of Veterans with PTSD attributed to their MST. Both treatments have shown initial promise, resulting in improvement in quality of life amongst female Veterans and positive long-term effects after completion of the treatment when compared to pharmacotherapy^5, 6^. However, approximately 33% of female Veterans drop out from such psychotherapeutic interventions, citing reasons such as limitations in access to care, reminders of the trauma, and overwhelmingly stigma despite mandated MST screenings in the VHA^6–8^.

Complementary and integrative health (CIH) interventions provide additional therapeutic support alongside traditional psychotherapeutic treatments in engaging Veterans who have experienced MST. CIH interventions such as mindfulness and yoga have increasingly been examined as treatments for MST and PTSD more broadly. Reviews of RCTs of CIH interventions for PTSD have found mindfulness interventions to outperform both active and passive controls in reducing military-related PTSD symptoms^9^, with yoga interventions being found to have similar, albeit more variable impacts on PTSD symptoms^10^. In the context of MST-related PTSD and female Veteran populations, it has been proposed that these interventions may be particularly well-suited due to their focus on bodily awareness and emotion regulation, as well their tendency to be administered in group formats which decrease social isolation and can increase feelings of empowerment^11^. In particular, a study investigating the effect of Restoration-Yoda Nidra Meditation (iRest) on PTSD and depressive symptoms in female Veterans with histories of MST, showed significant reductions in symptoms of both PTSD and depression, including negative thoughts of self-blame, over the course of treatment^12^. More recently, a first-of-its-kind RCT compared Trauma Center Trauma-Sensitive Yoga (TCTSY) to traditional CPT as a treatment for MST-related PTSD symptoms in a large cohort of female Veterans and found TCTSY to be equally effective in reducing PTSD symptoms, with moderate to high effect sizes^13^. Interestingly, although equivalent in symptom reduction, a significantly higher proportion of individuals in the TCTSY group completed treatment, indicating a greater degree of acceptability for TCTSY when compared to CPT^13^. This mirrors qualitative findings found in other studies, in which female Veterans have indicated high acceptability and even preferences for CIH interventions for MST-related symptoms^12, 14, 15^.

To our knowledge, although research findings have begun to show the effectiveness of mindfulness and yoga therapies as standalone CIH interventions, to our knowledge, lacking are studies that examine effectiveness of multimodal CIH interventions that target female Veterans at risk with histories of suicidal ideation or behavior that have experienced MST in improving their overall health and wellness. The present program evaluation firstly aims to investigate the potential impact of multifaceted CIH programming on symptoms of depression, anxiety, and PTSD, as well as perceived stress, pain, and sleep quality amongst female Veterans who have prior history of MST. Secondarily, it aims to investigate this CIH programming’s potential impact on these symptoms amongst these female Veterans who also have prior history of suicidal ideation or behavior aims to evaluate whether the multifaceted CIH programing leads to symptom improvements across these outcomes.

## METHODS

Measurement-based care was integrated into the RWC programing at inception. The RWC program evaluation was reviewed and determined by the Institutional Review Board of the VA hospital to be exempt from IRB review, and it was approved by the Quality Improvement Executive Committee. As such, for this program evaluation the authors utilized deidentified data from participant responses and did not perform informed consent, provide participants with monetary compensation, or register this program evaluation as a clinical trial.

### Resilience and Wellness Center program design

The Resilience and Wellness Center (RWC) at the Bronx VA is a program designed in October 2018 by a team of clinicians and researchers, focusing on CIH interventions that target key risk factors of suicidal behavior that have been described previously by our group, including details on specific programming offered^16^. Briefly, the program follows a cohort model in which Veterans participate in four weeks of various CIH treatment interventions and educational programming. These include meditation and mindfulness, yoga, physical exercise, nutrition, and motivational curriculum that equip participants with skills to improve their lifestyles and behaviors. Inclusion to RWC program is transdiagnostic and open to all Veterans receiving care across all services. Participants are clinician-referred and typically at-risk, vulnerable Veterans who are isolated, experiencing ongoing environmental stressors, and are interested in learning new coping strategies to make significant lifestyle changes. The majority of female Veterans in the present pilot study were referred by Women’s Health Primary Care clinic and were subsequently contacted by RWC staff to confirm understanding and willingness to attend a 5-days per week, 3-hour daily, 4-week program to be mindful to avoid scheduling conflicts while continuing their other treatments. RWC program engagement and outcome in female Veterans was evaluated for 15 independent cohorts participating from October 2018 to December 2019 (N=38 female participants).

### Measures

As part of the RWC programing aligned with patient-centered and measurement-based care^17, 18^ validated self-report instruments to assess mental and general health pre- and post-RWC program completion were integrated into the RWC for monitoring outcomes (described previously^16^). Of relevance to the present program evaluation, these included the Patient Health Questionnaire^19^ (PHQ-9) and Beck Depression Inventory II^20, 21^ (BDI) as measures of depressive symptoms, the Pittsburgh Sleep Quality Inventory^22^ (PSQI), and participant engagement as measured by daily program attendance as primary measures. In addition to these primary outcomes, participants also completed the Measure of Current Status^23^ (MoCS) to measure perceived coping ability, the Beck Anxiety Scale^24^ (BAS), PTSD Checklist for DSM-5^25^ (PCL), Perceived Stress Scale^26^ (PSS), and the Defense and Veterans Pain Rating Scale^27^ (DVPRS) as secondary measures. Data was collected at program inception for primary outcome measures (N=38), and for subset of participants (N=26) for secondary outcome measures.

### Data and statistical analyses

Analyses were performed using R 3.6.1^28^. based on 38 female participants in the RWC programing with history of MST. Demographic information was reported in Table 1 and compared between MST using t test for age and Fisher’s exact test for the categorical characteristics. Pre-post score differences were calculated (post - pre) and graphed by MST group for outlier inspection. Variables with outliers were either winsorized (i.e. censored to nearest non-outlier value), or non-parametric analysis was used. One-sided Wilcoxon rank sum tests were used when testing the MST vs No MST group difference for baseline scores, as well as pre-post score differences for the 9 clinical measurements of interest (namely PHQ-9, BDI, BHS, PCL, BAI, PSS, MoCS, DVPRS and PSQI), assuming participants in MST group had worse baseline scores and larger improvements compared to those in MST group. Moreover, ANCOVA were used to assess the MST group difference on pre-post score differences with adjustment for baseline scores. In a subset of 19 MST participants with prior history of suicide ideation or attempt, the effect size (measured as Cohen’s d) of pre-post score difference of the 9 clinical measurements were calculated with one outlier winsorized for score from the PSQI measure.

**Table 1.**
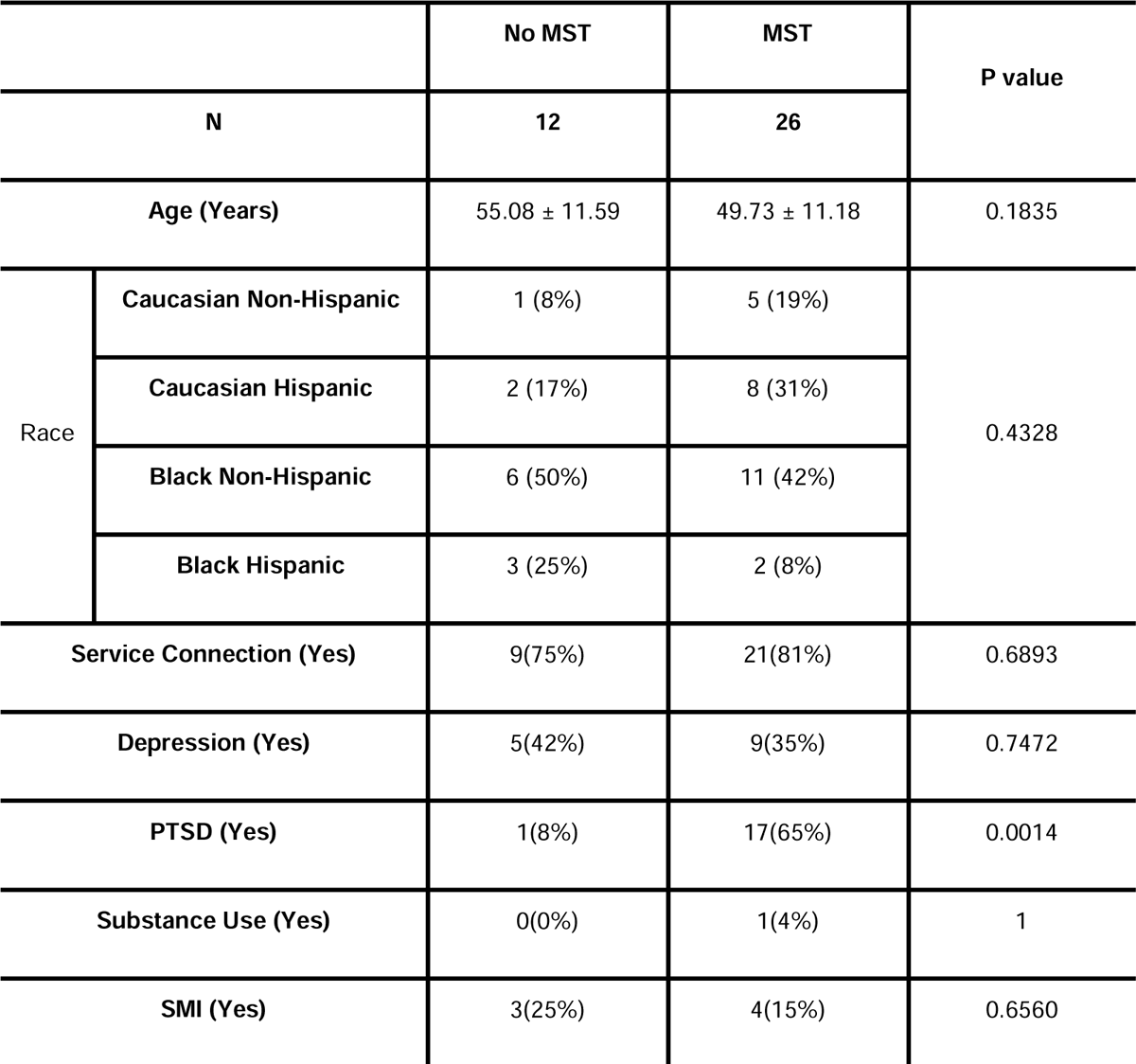
Demographics table reported by 38 female participants in the RWC program.

|  |  | No MST | MST | P value |
| --- | --- | --- | --- | --- |
| N |  | 12 | 26 |  |
| Age (Years) |  | 55.08 ± 11.59 | 49.73 ± 11.18 | 0.1835 |
| Race | Caucasian Non-Hispanic | 1 (8%) | 5 (19%) | 0.4328 |
|  | Caucasian Hispanic | 2 (17%) | 8 (31%) |  |
|  | Black Non-Hispanic | 6 (50%) | 11 (42%) |  |
|  | Black Hispanic | 3 (25%) | 2 (8%) |  |
| Service Connection (Yes) |  | 9(75%) | 21(81%) | 0.6893 |
| Depression (Yes) |  | 5(42%) | 9(35%) | 0.7472 |
| PTSD (Yes) |  | 1(8%) | 17(65%) | 0.0014 |
| Substance Use (Yes) |  | 0(0%) | 1(4%) | 1 |
| SMI (Yes) |  | 3(25%) | 4(15%) | 0.6560 |

## RESULTS

Among these 38 female participants, 26 (68%) had a positive MST screen in their electronic medical record (EMR) for military sexual trauma during service. Demographic characteristics (Shown in Table 1) did not differ between MST and no MST participants in general, except for PTSD. PTSD prevalence differed significantly between the MST and No MST groups (p = 0.0014), with 65% of MST participants having a PTSD diagnosis in their EMR in comparison to only 8% of No MST participants. Assessment of mental and physical health symptoms performed both at baseline and after the completion of the RWC program, showed that participants who endorsed MST experience reported significantly worse baseline scores in 5 out of 9 measures of interest compared to participants without MST, including depressive symptoms (measured by PHQ-9 and BDI scores; p = 0.0402 and 0.0360 respectively), PTSD symptoms (measured by PCL; p =0.0254), perceived stress (measured by PSS, p = 0.0121), and sleep quality (measured by PSQI; p= 0.0208) shown in Table 2. We further examined whether the MST group showed greater improvement after program completion as measured by these symptom domains. In comparison to the no MST group, significantly larger improvements (pre-post score difference, delta = post – pre) for the MST group were found for PHQ-9, BDI and PSS (p = 0.0156, 0.0211 and 0.0351 respectively, Table 3); However, when adjusting for baseline scores, the group differences were no longer significant for these 3 measures (p = 0.3120, 0.1181 and 0.8840 for PHQ-9, BDI and PSS correspondingly). Further, we explored if there were improvements in clinical measurements among female Veterans endorsing MST who had prior history of suicide ideation or attempt. Among the subset of 19 MST participants with suicide ideation or attempt history, median to large RWC treatment effects were observed for all symptom measures (Table 4).

**Table 2.**
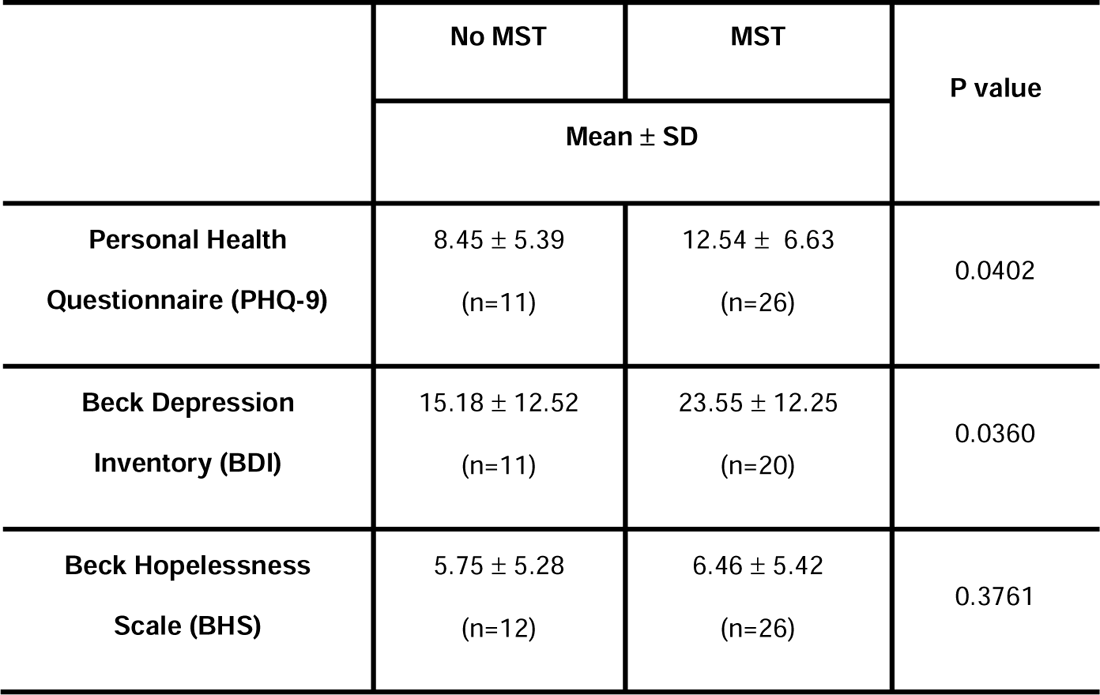
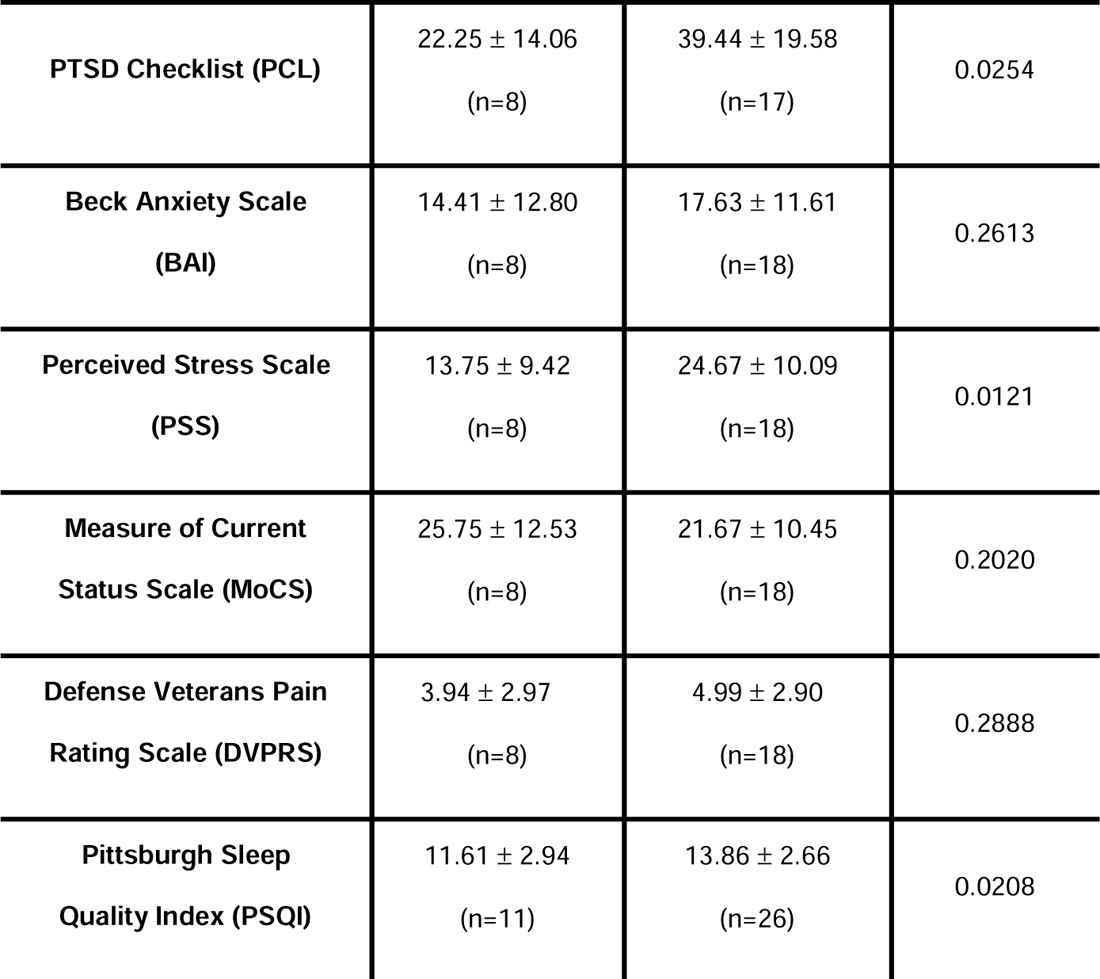
Statistics of MST group comparison in baseline scores. Mean, standard deviation, and sample size of each clinical measurement are reported by MST group and P values come from one-sided Wilcoxon rank sum tests.

**Table 3.**
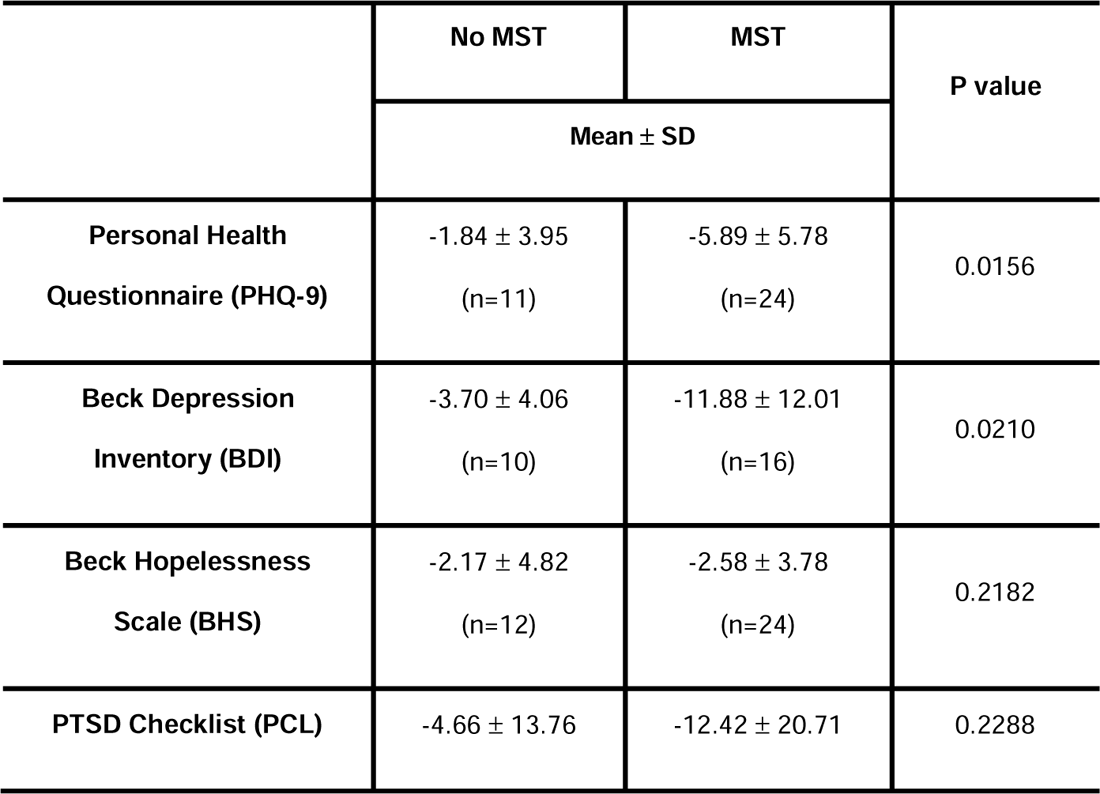
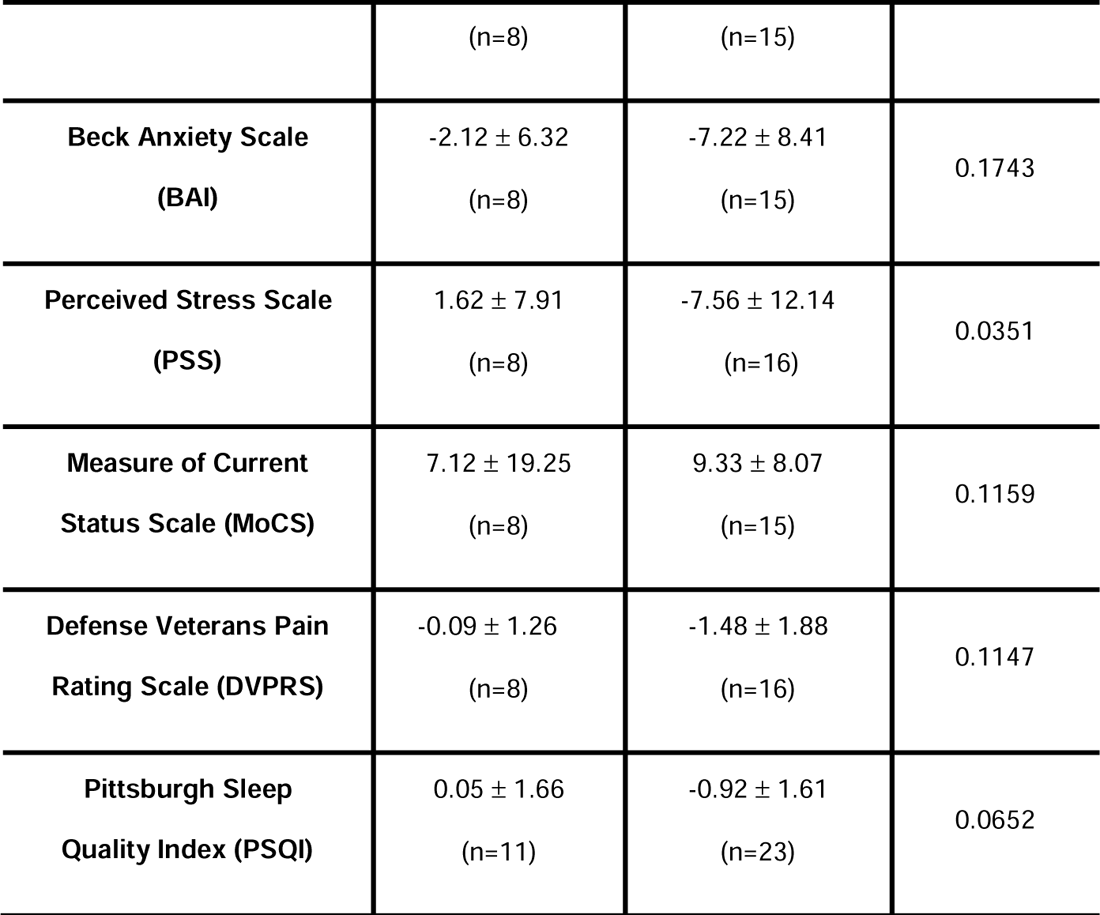
Statistics of MST group comparison in pre-post score difference (post – pre). Mean, standard deviation, and sample size of each clinical measurement are reported by MST group and P values come from one-sided Wilcoxon rank sum tests.

**Table 4.**
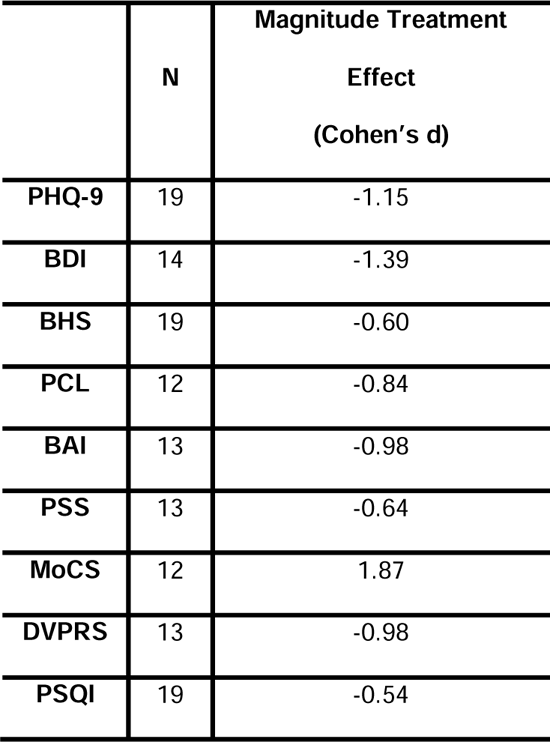
Significance of treatment outcomes as measured by associated effect sizes (Cohen’s d) in MST participants who had a history of suicide attempt. Values of d, correspond to negligible (−0.2<d<0.2), small (0.2≤d<0.5,-0.5<d≤-0.2), medium (0.5≤d<0.8;-0.8<d≤-0.5), and large effect size beyond.

|  | N | Magnitude Treatment<br>Effect<br>(Cohen's d) |
| --- | --- | --- |
| **PHQ-9** | 19 | -1.15 |
| **BDI** | 14 | -1.39 |
| **BHS** | 19 | -0.60 |
| **PCL** | 12 | -0.84 |
| **BAI** | 13 | -0.98 |
| **PSS** | 13 | -0.64 |
| **MoCS** | 12 | 1.87 |
| **DVPRS** | 13 | -0.98 |
| **PSQI** | 19 | -0.54 |

## DISCUSSION

Although individual CIH interventions such as mindfulness meditation and trauma-informed yoga have been found to be effective in reducing mental health symptoms associated with MST in female Veterans, to our knowledge, no research has yet examined the impact of an intervention that combines multiple CIH treatments into a holistic treatment framework. In light of this, the present study aimed to evaluate the potential psychological benefits of such an integrated CIH intervention on female Veterans who experienced MST. Significantly greater reductions in symptoms of depression and stress were found in the MST group in comparison to those without MST over the course of the multimodal RWC programming delivered in this study. Veterans who experienced MST in general had worse baseline scores for depressive and PTSD symptoms, perceived stress, and sleep quality, which when adjusted for in the analyses explained away these observed group differences in symptom reductions. Notably, there was an overall improvement in a wide range of mental and general health symptoms in the at-risk MST participants with prior history of suicidal ideation or attempt. Together, these results indicate that the multimodal CIH intervention administered in this study may have been effective in reducing the severity of symptoms related to MST.

The results presented here also add to the growing body of research findings demonstrating CIH interventions to be effective in attenuating mental health symptoms, specifically PTSD symptoms stemming from MST^9, 10^. CIH interventions are suggested to be particularly well-suited for individuals with these symptoms for several reasons. First, use of mindfulness and meditation techniques often found in evidence-based treatments may help increase individuals’ awareness of being present when experiencing stressful symptoms, leading to greater acceptance^11, 12^. The integration of physical movement and breathing techniques, particularly in yoga, may also increase body interoception and improve body image, both of particular importance to survivors of MST whose trauma is often experienced primarily in the body^13^. The focus of these interventions on breathing and the body may further impact PTSD symptoms such as sleep disturbances by fostering more adaptive physical stress responses by regulating sympathetic and parasympathetic nervous system activation^29^. Such a pathway is corroborated by work that has found decreases in Veterans’ PTSD symptoms during CIH interventions to be linked to corresponding decreases in salivary cortisol^30^. Finally, amongst women Veterans with MST, the delivery of CIH interventions often in a group format, can send a message to Veterans who participate that they are not alone, reducing symptoms of social isolation and self-stigma^11^, which may also contribute to the high degree of acceptability for these interventions in this group. Our findings demonstrate for the first time the potential positive impact of a cohort-based multimodal CIH intervention amongst at-risk female Veterans in a likewise fashion to individual CIH interventions.

This program evaluation study has several limitations including the lack of objective measures, control group, and longitudinal follow up data collection. As all of the measures were self-report, the programming would have benefited from inclusion of clinician-administered measures as well as physiological measures including cortisol levels^31, 32^ or HRV^33, 34^ associated with stress and depressed mood. Another significant limitation is the lack of a control group, since this was a quality improvement evaluation, which does not allow us to conclude study efficacy regarding whether the present CIH interventions were the cause of observed symptom reductions.

Similarly, since the programming was based off of monthly cohorts, assessing participants at the beginning and at the end of the month, there were no longitudinal follow-up measures to assess long-term symptom trajectories in MST survivors after conclusion of the CIH interventions. Research studies building on the findings from this program evaluation can include a control group and follow-up measures in their design, which would allow for stronger conclusions regarding whether multimodal CIH interventions can directly lead to MST symptom improvement, as well as how long symptom improvement may last following cessation of the intervention.

Future CIH interventions for Veterans with MST could expand upon this work in several ways. While our current programming was offered to our Veterans was completely in-person, future programming could offer CIH interventions through virtual platforms, as reported previously by this group^35^, which could substantially broaden access to care for Veterans with history of MST. Investigating the potential benefits of CIH modalities virtually amongst Veterans with MST is necessary to continue access to care, especially to Veterans who may have difficulties coming in-person to receive care due to barriers relating to transportation issues and severe physical and mental health challenges. Future iterations of these interventions could also explore their effectiveness in currently understudied populations such as male and LGBTQ+ Veterans with histories of MST. Fewer investigations of male MST exist due to male MST encounters often not being disclosed because of the concern for severe stigmatization, often leading to worsening mental health issues for which such CIH interventions may be an effective treatment^36^. The effectiveness of CIH interventions in treating LGBTQ+ Veterans with histories of MST is similarly understudied and holds particular promise due to this populations’ higher rates of MST-related mental illness^37, 38^ and apparent preference for CIH interventions^39^ when compared with their non-sexual minority counterparts. In sum, Complementary and Integrative Health interventions show great potential in improving the overall wellness of Veterans with MST, providing a pathway to promote inclusivity and broaden therapeutic efforts.

## Data Availability

Deidentified data supporting the conclusions of this article can be made available by the authors, on condition that intuitional and ethical requirements for sharing data are met.

## Acknowledgements

The authors would like to thank the Veterans for their involvement in this quality improvement project and the Resilience and Wellness program, as well as the clinicians and instructors for contributing their time and expertise. They would also like to thank the David Lynch Foundation’s Trauma Project for contributing their TM teachings as well as Dr. Nicholas Bowersox, Director, QUERI Center for Evaluation and Implementation Resources (CEIR), for his guidance and support in carrying out this Quality Improvement project.

